# Mechanical thrombectomy of the anterior circulation ischemic stroke whatever the low-volume operator : comparison to a 90 days follow-up

**DOI:** 10.1101/2022.10.13.22280979

**Authors:** Guillaume Herpe, Victor Dumas, Rémy Guillevin

**Affiliations:** University Hospital Centre Poitiers, Radiology, Poitiers, Vienne, 86000, FR; Université de Poitiers Laboratoire de Mathématiques et Applications, DACTIM-MIS, Chasseneuil, Nouvelle Aquitaine, FR

## Abstract

**Background:** To date, benefit of successful recanalization of acute stroke has been described mainly for high volume operator.

**Purpose:** To evaluate if a successful MT performed by a low-volume operator is a strong predictor of patient’s good clinical outcome.

**Materials and Methods:** A consecutive monocentric observational study from June 1, 2016 to December 31, 2018 at the University Hospital of Poitiers. Patient management included mechanical thrombectomy and eventual IVT. The interventional neuroradiologist on duty classified the recanalization according a standardized mTICI reading grid. ∆mRS was defined as the difference between 90 days and initial mRS. Association were studied using Chi-square test and Pearson’s and Fisher’s correlation with a threshold at 0.05. The main criterion was the reliability between mTICI and ∆mRS.

**Results:** Among the 304 MT, 127 patients were finally included. High mTICI score was significantly associated with high ∆mRS (p = 0.04), using the mTICI dichotomization, patients with successful MT had significantly better outcome than patient with lower mTICI whatever the initial mRS (p = 0.008). There was a correlation between age categories greater than or equal to 80 y.o (p=0.03) and high initial NIHSS categories greater than or equal to 18 (p=0.004) with regard to the ∆mRS. No statistical significance were obtained between sexe (p=0.63), diabetes mellitus (p=0.25), VIT (p=0.16) and thrombus’s lateralization (p=0.26) with regard to ∆mRS.

**Conclusion:** This study highlights the place and performances of low-volume operator in the treatment of acute stroke.

**Summary Statement:** Despite low-volume operators, a successful mechanical thrombectomy is a strong prognostic factor for good clinical outcome.

**Key Results:**

- Mechanical thrombectomy performed in a low volume center is a safe therapy and strongly linked to good clinical outcome.
- Our low volume-operators centre has reached a high reperfusion rate and a majority of good clinical outcome
- This study highlights the place and performances of low-volume operator in the treatment of acute stroke.

## mTICI score as an independant prognosis factor after mechanical thrombectomy of the anterior circulation in a low volume center : comparison to a 90 days follow-up

Acute stroke, is the first cause of acquired disability in adults, the second leading cause of dementia and the third leading cause of death in the world. The annual prevalence of stroke is up to 6/1,000 subjects. The increasing number of stroke led to a public health issue, for which the most effective, standardized, and consistent care must be offered.

80% of the strokes are caused by cerebral ischemia (CI), as a result of the interruption of cerebral blood flow (CBF). The CBF impairment due to a decrease in cerebral perfusion, falling below the threshold of physiological self-regulation of cerebral blood flow, thus leading to cerebral hypoxemia. As a result, phenomena quickly lead to “neuronal silence”, also called the penumbra zone, then to irreversible necrosis, i.e. cerebral infarction, by neuronal death via depolarization mechanisms, oxidative stress and excitotoxicity (1).

The therapeutic strategy for acute stroke must be initiated urgently and is based on intravenous (IV) thrombolysis with rt-PA associated with Mechanical Thrombectomy (MT) (2). The superiority of the combination of IVT and MT versus IVT alone was demonstrated by Berkhemer and al (3).

Numerous studies, including the meta-analysis of Rha and Saver (4), agreed in demonstrating that successful reperfusion was a powerful predictor of the patient’s good clinical outcome and of lower mortality. These studies were focused on large population and/or high volume center. Since the mechanical thrombectomy is a highly technical procedure, whether it should be performed in low volume center remains debated as the experiment of the operators working in theses centers are supposed to be lower.

The society of Vascular and Interventional Neurology (SVIN) established Stroke Interventional Laboratory Consensus (SILC) criteria to develop stroke interventional laboratory in the era of stroke thrombectomy for large vessel occlusions. They concluded that a low-volume operator, <50 numbers/year, should only work in high volume laboratory >100 numbers/year (5), which corresponds to the practice of our center.

The aim of our study was to evaluate if a successful MT performed by a low-volume operator is a strong predictor of patient’s good clinical outcome.

## MATERIAL AND METHOD

We have conducted a retrospective consecutive monocentric observational study from June 1, 2016 to December 31, 2018 at the University Hospital of Poitiers.

All consecutive patients who underwent mechanical thrombectomy were eligible.

Patients were recorded, without age limit. They underwent a neurological examination by a senior neurologist with more than 5 year’s experience in stroke.

Patient management included mechanical thrombectomy and eventual Intra Veinous Thrombolysis (IVT). Management was discussed on a case-by-case basis after consensus between vascular neurologists and interventional neuroradiologists.

The following clinical data were recorded from the patient electronic file : sexe, diabetes mellitus, thrombus location and lateralization, National Institutes of Health Stroke Scale (NIHSS) score at pretreatment, use of IVT, modified Thrombolysis in Cerebral Infarction (mTICI), number of passes, mRS at pretreatment and at day 90 (mRS 90). Age of the patient were also recorded and dichotomized into : greater or equal than 80 years old (y.o), or less than 80 y.o. NIHSS score was also dichotomized into less or equal than 17 and greater than 17. Theses parameters consisted in the previously published independent risk factors for poor clinical outcome despite a high mTICI score (6) (7)(8) (9) (10) (11).

Exclusion criteria were patients with thrombus other than M1 or M2, insufficient DSA for technical reasons, patients who were lost to follow-up and/or whose modified Rankin score at day 90 (mRS 90) was not assessable.

Informed consent was obtained, and study was approved by the local ethic committee.

All patients underwent a mechanical thrombectomy in the angiography room of the Poitiers University Hospital, using the same technic. The technic along with the acquisition parameters are provided within the supplementary material (S1).

### Image reading

During the procedure, cerebral angiographies were read blindly from our study by the interventional neuroradiologist on duty (list of interventional neuroradiologist, stating years of experience and number of MT performed during this stydy is provided within supplementary material S2). Reading was performed using standardized windowing on a dedicated reading software.

The interventional neuroradiologist on duty classified the patient according a standardized mTICI reading grid. mTICI reading grid is provided within the supplementary material (S3).

mTICI score were secondly dichotomized into successful procedure (near complete reperfusion : mTICI 2c or complete reperfusion, mTICI 3) or unsuccessful procedure (mTICI 2b and under).

### Reference standard

All analyzed patients underwent a neurological examination by a senior neurologist 90 days after the stroke. The examination included at least a modified Rankin Score blinded from the results of the mechanical thrombectomy. mRS grid is provided within the supplementary material (S4).

Both the initial and 90 days mRS scores were recorded. Delta mRS (∆mRS) was defined as the difference between 90 days and initial mRS.

The clinical evolution was dichotomized into favorable functional evolution as a ∆mRS score less than or equal to 2 and poor clinical outcome in case of a score over than 2.

## Statistical analysis

Patient characteristics were presented using mean and standard deviation (SD) for quantitative variables and proportions (%) for categorical variables. Tests for significant difference between patients with good (mRS_≤_2) and poor (mRS>2) clinical outcome was assessed by Pearson’s chi-square test and Fisher’s exact test for categorical variables. The main criterion was the reliability between mTICI and ∆mRS.

The calculation of the patient’s good clinical outcome at 3 months was calculated from stepwise logistic regression. Variables significantly related in the univariate analysis were selected in the model.. The model uses the Rankin category at 90 days as the variable to be explained ; and age, baseline NIHSS score and mTICI score dichotomously (0-2b versus 2c-3) as the explanatory variable. The goodness-of-fit of this model was estimated using the Hosmer and Lemeshow chi-square test with a level of p _≤_ 0.05. The best model is obtained by taking the three parameters.

## RESULTS

### Demographics

304 patients underwent a MT during the time of the study consisting in 121.6 MT per year, and 20.27 MT per operator. Of these 304 patients, 81 were excluded secondarily because they had a thrombus with a location other than anterior circulation, 10 because mTICI score was not assessable due to motion artifacts and 86 because their 90-day SRM score was not assessable (lost to follow-up). A total of 127 patients were thus included.

The flow chart is provided in Figure 1.

**Figure 1:**
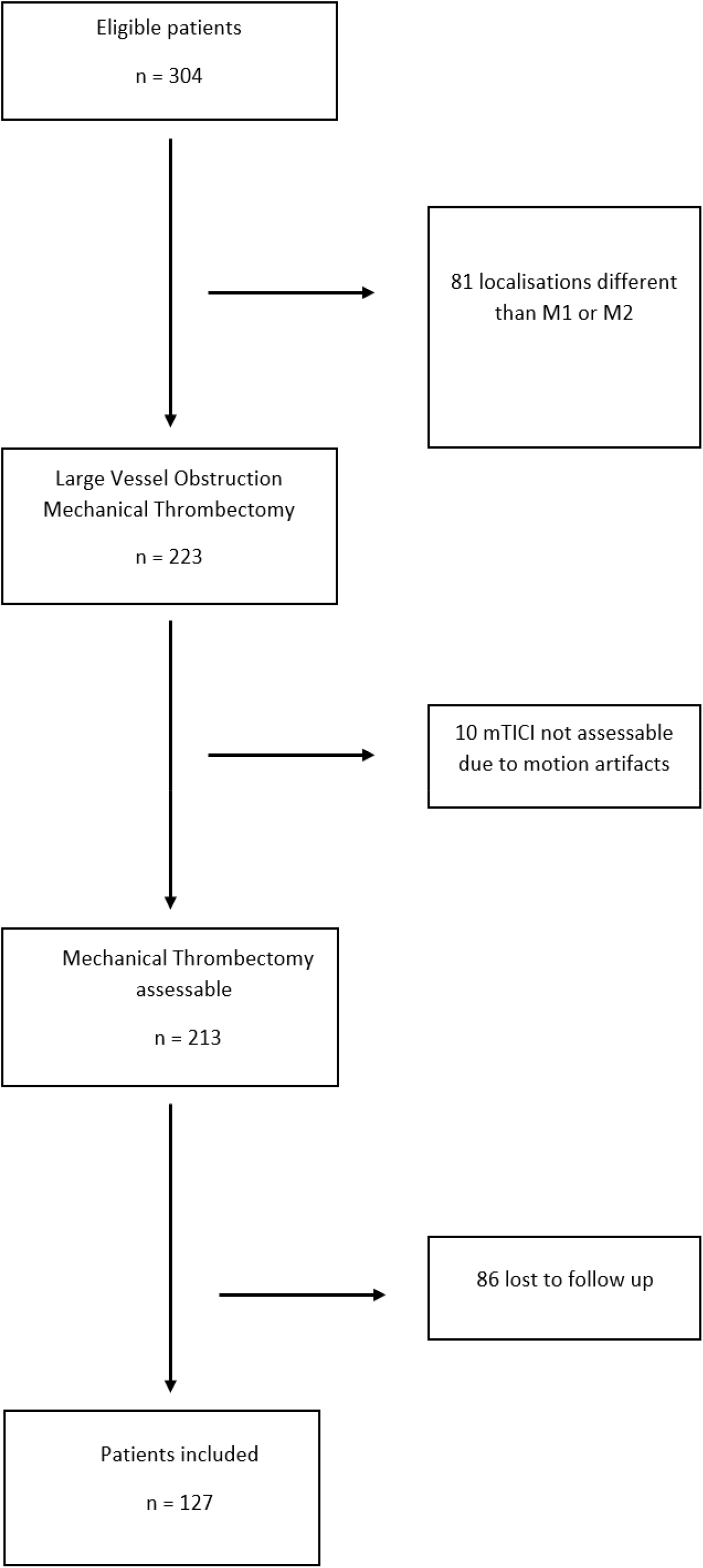
Flow Chart of the study patient.

The mean age (± standard deviation) of the included patients was 70 y.o (± 17.2). Regarding the substantial demographics, 37 patients were over 80 years old (29% 37/127), 19 were diabetic (15% 19/127).

The mean pretreatment NIHSS score was 15,7 (± 5). 78 patients received IVT(62.4% 78/127).

The mean number of passes was 2.1 (±1.6). Thrombus were in the left anterior circulation in 51.9 % (66/127) and in the right circulation in 48% (61/127).

5 MT (3.9 % 5/127) were evaluated mTICI0, 10 (7.9% 10/127) mTICI 2a, 39 (30.7% 39/127) mTICI 2b, 12 (9.5% 12/127) mTICI 2c and 61 (48.0% 61/127) mTICI 3. None of the patient had a mTICI score of 1.

Mean ∆mRS was 3.1 (±2.4). 72 patients 56.7% 72/127) had a ∆mRS less or equal than 2 ; 55 (53.4% 55/127) greater than 2 Demographic and MT characteristics are summarized in Table 1.

**Table 1.** illustrates the demographics of the overall population patients and statistical differences within the sub groups. Data in parentheses are percentages with 95% confidence interval. Standard deviation are calculated with 95% interval.

|  | Overall | Male | Female | Test | P value |
| --- | --- | --- | --- | --- | --- |
| Mean age y.o (SD) | 70 (17.2) | 69.3 (18.6) | 71.2 (15.9) | Chi2 | 0.24 |
| 0-79 | 90 (71%) | 44 (71%) | 46 (71%) | Chi2 | 1.00 |
| ≥80 | 37 (29%) | 18 (29%) | 19 (29%) |  |  |
| Number of passes (SD) | 2.1 (1.6) | 2.2 (1.7) | 2 (1.5) | Chi2 | 0.98 |
| Diabetes mellitus |  |  |  |  |  |
| No | 108 (85%) | 51 (82.3%) | 57 (87.7%) | Chi2 | 0.54 |
| Yes | 19 (15%) | 11 (17.7 %) | 8 (12.3 %) |  |  |
| Baseline NIHSS (SD) | 15.7 (5) | 15.3(5.2) | 16 (4.8) | Chi2 | 0.62 |
| TIV |  |  |  |  |  |
| No | 49 (37.6%) | 25 (40.4%) | 24 (37%) | Chi2 | 0.83 |
| Yes | 78 (62.4%) | 37 (59.6%) | 41 (63%) |  |  |
| mTICI |  |  |  |  |  |
| 0 | 5 (3.9%) | 3 (4.8%) | 2 (3.1%) | Fisher | 0.80 |
| 1 | 0 (0%) | 0 (0%) | 0 (0%) |  |  |
| 2a | 10 (7.9%) | 5 (8.1%) | 5 (7.7%) |  |  |
| 2b | 39 (30.7%) | 16 (25.8%) | 23 (35.4%) |  |  |
| 2c | 12 (9.5%) | 7 (11.3%) | 5 (7.7%) |  |  |
| 3 | 61 (48.0%) | 31 (50%) | 30 (46.1%) |  |  |
| mRS at 90 days (SD) | 3.1 (2.4) | 3.1 (2.5) | 3.1 (2.3) | Chi2 | 0.21 |
| ΔmRS (SD) | 2.7 (2.3) | 2.7 (2.3) | 2.7 (2.3) | Chi2 | 0.47 |

## Statistical

On univariate analysis, a high mTICI score was significantly associated with high ∆mRS (p = 0.04).

Using the mTICI dichotomization, the patient with successful MT (near complete to complete reperfusion) had significantly better outcome than patient with lower mTICI whatever the initial mRS (p = 0.008).

The number of passes was significantly higher in the subgroup mTICI 2a (p=0.003) than in the others groups. No statistical significance was reached between number of passes (p=0.17) and ∆mRS.

Regarding the usually described independant risk factors, we found correlation between age categories greater than or equal to 80 y.o (p=0.03) and high initial NIHSS categories greater than or equal to 18 (p=0.004) with regard to the ∆mRS.

Regarding the region of stroke (Left or Right). A Pearson’s chi-squared test failed to show an association between region and mTICI score (p=0.384) or between region and Rankin change (p=0.257). No statistical significance were obtained between sexe (p=0.63), diabetes mellitus (p=0.25), VIT (p=0.16) and thrombus’s lateralization (p=0.26) with regard to ∆mRS.

Table 2 illustrates the main correlation within the identified risk factors.

**Table 2.** illustrates the main correlation within the identified risk factors along with the statistical test used to assess theses correlations.

| | ΔmRS 0-2 | ΔmRS 3-6 | Test | P value |
| --- | --- | --- | --- | --- |
| **Patients risk factors** |  |  |  |  |
| Overall | 72 (56.7%) | 55 (53.4%) | Chi2 | 0.63 |
| Sexe |  |  |  |  |
| Male | 37 (51%) | 25 (45%) | Chi2 | 0.62 |
| Female | 35 (49%) | 30 (55%) |  |  |
| Diabete mellitus |  |  |  |  |
| No | 64 (89%) | 44 (80%) | Chi2 | 0.25 |
| Yes | 8 (11%) | 11 (20%) |  |  |
| Age |  |  |  |  |
| 0-79 | 57 (79%) | 33 (60%) | Chi2 | 0.03 |
| $\geq 80$ | 15 (21%) | 22 (40%) | | |
| VIT |  |  |  |  |
| No | 22 (31%) | 25 (45%) | Chi2 | 0.16 |
| Yes | 48 (69%) | 30 (55%) |  |  |
| Baseline NIHSS |  |  |  |  |
| 0-17 | 51 (71%) | 24 (44%) | Chi2 | 0.004 |
| 18-25 | 21 (29%) | 31 (56%) |  |  |
| Number of passes |  |  |  |  |
| 1 | 45 (63%) | 22 (40%) | Fisher | 0.17 |
| 2 | 11 (16%) | 13 (24%) |  |  |
| 3 | 6 (9%) | 6 (11%) |  |  |
| 4 | 3 (4%) | 6 (11%) |  |  |
| 5 | 4 (6%) | 3 (5%) |  |  |
| 6 | 1 (1%) | 3 (5%) |  |  |
| 7 | 1 (1%) | 2 (4%) |  |  |
| mTICI |  |  |  |  |
| 0 | 1 (1%) | 4 (7%) | Fisher | 0.04 |
| 1 | 0 (0%) | 0 (0%) |  |  |
| 2a | 6 (8%) | 4 (7%) |  |  |
| 2b | 16 (22%) | 23 (42%) |  |  |
| 2c | 9 (13%) | 3 (6%) |  |  |
| 3 | 40 (56%) | 21 (38%) |  |  |
| $\geq 2c$ | 49 (39%) | 23 (18%) | Fisher | 0.0008 |

## DISCUSSION

This study is to our knowledge one of the first studying the feasibility of mechanical thrombectomy in a low volume center. We demonstrated and confirmed that reperfusion rate (mTICI) was a predictor of good

clinical outcome indenpendantly of the others identified risks. Theses results in a small volume center are in agreement with the previously published studies. The MERCI trials found association with good outcome (p<0.0018) (6). In the penumbra stroke trial, good outcome occurred in 29% of the recanalized patients versus 9% of the non-recanalized patients (p=0.06) (12).

The mechanical reperfusion therapeutics seems to be one of the only independent prognostic factor that can be triggered by the institution confirming the key role of mechanical thrombectomy.

A greater number of passage was statistically related to mTICI 2a final reperfusion (p=0.003). This can be explained by the fact that when the procedure is presented as a complicated but feasible procedure from the onset, the interventional neuroradiologist is tempted to insist and therefore to multiply the number of passages. On the contrary, when the procedure appears to be complicated with little hope of effective removal of the obstacle, the operator prefers not to multiply the passages in order to avoid possible complications.

Younger age, and lower NIHSS score were also directly associated with good outcome, which had already been found to be independent factors of good clinical outcome (6) (7) (8) (9).

It’s particularly important to assess the impact of age on post treatment outcome since the ederly represent a growing proportion of acute stroke sufferers. The shift analysis of the meta HERMES analysis found a significant benefit from the combined treatment with the population over 80 years of age, with more than half of the patients in this age group being over 80 years of age achieving functional autonomy at 3 months (13). Loh Y and al had also demonstrated that elderly patients with an acceptable initial neurological status could benefit from mechanical thrombectomy (14).

A good clinical outcome was achieved for 72% patients in our study (Table1), whereas a previously published meta-analysis assessed 46% of patients with an mRS score 0–2 at 90 days in the endovascular thrombectomy population (15).This could be explained by our inclusion criteria. We selected only anterior circulation stroke as it is the most frequent and standardized procedures. Since anterior circulation stroke are of better prognostic than posterior circulation, the results of our clinical outcome are finally in agreement with the literature.

However, some limitation have to be considered.

We did not collected all the described poor prognostic factors such as the delay between the first signs and the start of thrombectomy, high blood pressure, collaterality network, thrombus density. As it was a retrospective study, some few data were available for theses parameters.

We did not study the onset-to-door-time for instance whereas it is a strong factor for prognosis. All the included patients had a less than 6 hours onset-to-door-time and 86 patients (39.8%) of the eligible population were lost to follow-up, this is a high rate but it could be explained by the large geographical area of our reference center.

## CONCLUSION

We demonstrated that successful MT was a strong prognostic factor for good clinical outcome even in a center with low volume operator.

## Data Availability

All data produced in the present work are contained in the manuscript

## Abbreviations

MT: Mechanical Thrombectomy
IVT: Intra Veinous Therapy
mTICI: Modified Thrombolysis In Cerebral Infarction
mRS: Modified Rankin Score

## SUPPLE MENTARY MATERIAL

**S1 : <u>Modified Rankin score (mRS) :</u>**

- No symptoms.
- No significant disability. Able to carry out all usual activities, despite some symptoms.
- Slight disability. Able to look after own affairs without assistance, but unable to carry out all previous activities.
- Moderate disability. Requires some help, but able to walk unassisted.
- Moderately severe disability. Unable to attend to own bodily needs without assistance, and unable to walk unassisted.
- Severe disability. Requires constant nursing care and attention, bedridden, incontinent.
- Dead.

**S2 : <u>List of interventionnal neuroradiologist, years of experience and number of mechanical thrombectomy (MT) performed during this study</u>:**

S.V., 15 years of experience, 40 MT ; S.B., 10 years of experience, 27 MT; P.C., 8 years of experience 21 MT; G.V., 7 years of experience 22 MT ; G.H., 6 years of experience 10 MT ; A.G., 4 years of experience, 7 MT.

**S3 : <u>Technic along with the angiogram’s acquisition parameters :</u>**

A Newton TERUMO catheter (n°RFEH15010M), 5 French, was introduced through the femoral artery to the internal carotid artery, and the radiological study was performed by conventional antero-posterior view and profile (SIEMENS Axium biplane sensor - Artis (Germany). The acquisition parameters were standardized a voltage of 76kV and an amperage of 120mA, collimation of 4.8 cm, and a rate of 3 images per second). The contrast agent (Iomeron 300mg/ml, Iomepro, BRACCO, Italy), was injected into the internal carotid artery at a rate of 4 cc/second, at a dose of 8 ml, using a MEDRAD automatic injector (Mark V pro VIS, Bayer Healthcare, USA).

Digital subtraction was performed for the angiographic study using SIEMENS reconstruction software.

The devices used during the procedure were not evaluated and depended on the interventionnal neuroradiologist. Most of th preocedures were performed using a stent retrieving technics.

An X-Ray acquisition was performed before and after mechanical thrombectomy. Dynamic X-rays acquisitions were performed over time: 2 images per sec during 20 seconds after intra-arterial injection resulting in a set of 40 images from arterial phase, parenchymal phase and then venous phase. The data were then extracted using Maincare PACS station in DICOM files and then processed using the algorithms.

**S4 : <u>Modified thrombolysis in cerebral infarction score (mTICI) :</u>**

mTICI 0 : No recanalization.

mTICI 1 : Minimal recanalization with limited distal vascularization.

mTICI 2a : Partial recanalization, with distal vascularization less than half of the half of the occluded vascular territory.

mTICI 2b: Partial recanalization, with distal vascularization of more than half of the occluded half of the occluded vascular territory.

mTICI 2c : Almost complete recanalization with slow flow in some distal branches or or distal emboli.

mTICI 3 : Complete recanalization of the entire vascular territory.

## Notes

### Competing Interest Statement

The authors have declared no competing interest.

### Funding Statement

This study did not receive any funding

### Author Declarations

Ethics committee/IRB of Chu de Poitiers gave ethical approval for this work

